# Racial and Ethnic Disparities in Statin Adherence: Insights from the All of Us Research Program

**DOI:** 10.1101/2025.08.26.25334490

**Authors:** Gabriela Escobar, Zahra Azizi, Anne de Hond, Ashley Adanna Lewis, Madelena Y. Ng, Fatima Rodriguez, Tina Hernandez-Boussard

**Affiliations:** Stanford University, Stanford, CA, USA; Division of Cardiovascular Medicine, Stanford University, Stanford, CA, USA; Julius Center for Health Sciences and Primary Care, University Medical Center Utrecht, Utrecht, the Netherlands; Department of Biomedical Data Science, Stanford University, Stanford, CA, USA; Center for Digital Health, Stanford University, Stanford, CA, USA

**Keywords:** Social Determinants of Health (SDoH), Statin Adherence, Coronary artery disease, Health Disparities, Race and Ethnicity, All of Us cohort

## Abstract

**Background:** Statin adherence impacts cardiovascular outcomes, yet disparities persist. Understanding sociodemographic factors and barriers is crucial for targeted interventions.

**Objective:** To investigate the relationship between sociodemographic factors and statin adherence across racial and ethnic groups.

**Design:** This retrospective study examined sociodemographic factors, prescription records, clinical factors, and responses from the Demographic, Drug Exposure, Healthcare Utilization Survey (HUS) in the All of Us (AoU) cohort. Multivariable logistic regression models assessed the impact of sociodemographic factors on adherence stratified by race.

**Participants:** Adult participants with statin prescription records. Subgroup analyses included those who responded to the HUS.

**Exposures:** Statin prescription

**Main Outcome(s) and Measure(s):** Percent days covered (PDC), calculated as the proportion of days within a year in which a person prescribed a statin filled a prescription. Adequate adherence was defined as PDC ≥ 80%.

**Results:** Of the 17,029 participants with a statin prescription, the mean statin PDC was 57%, with 66% reporting a PDC ≤ 80%. Racial and ethnic differences in adherence were observed, with Non-Hispanic White (NHW) participants having a median PDC of 74% (IQR [0.25,0.98]), Non-Hispanic Black (NHB) 49% (IQR [0.25,0.98]), and Hispanic participants 25% (IQR [0.08,0.49]). NHW participants faced employment barriers (OR 0.63, 95% CI [0.46, 0.86]) and provider inaccessibility (OR 0.56, 95% CI [0.40, 0.76]) as significant factors for lower adherence. NHB participants experienced patient anxiety (OR 0.53, 95% CI [0.30, 0.90]) and financial barriers (OR 0.65, 95% CI [0.50, 0.85]), while Hispanic participants showed patient anxiety (OR 0.14, 95% CI [0.02, 0.60]) and immigrant status (OR 0.36, 95% CI [0.17, 0.76]) as significant factors for lower adherence.

**Conclusions and Relevance:** To address cardiovascular disease disparities, it is crucial to recognize unique sociodemographic barriers to statin adherence within racial and ethnic groups. Our findings highlight the need for tailored strategies considering the diverse barriers each group faces. Targeted interventions can bridge adherence gaps and improve cardiovascular outcomes across populations. This approach recognizes that while race and ethnicity may correlate with specific barriers, it is the underlying SDoH that often play a pivotal role in statin adherence.

**Key Points:** *Question:* What are predictors and barriers to statin adherence in the All of Us cohort?

*Findings:* Among 17,029 statin users, 57% reported low statin adherence, which varied by racial and ethnic groups and was associated with social determinants of health (SDoH). For Non-Hispanic Whites, employment barriers and provider inaccessibility were linked to lower adherence. Non-Hispanic Blacks faced patient anxiety and financial barriers, while Hispanics’ adherence was impacted by immigrant status, healthcare coverage and patient anxiety.

*Conclusions:* Unique SDoH influence statin adherence among diverse populations. Tailored interventions addressing specific barriers are needed to improve adherence and reduce cardiovascular disease risk equitably and effectively.

## Introduction

Atherosclerotic cardiovascular disease (ASCVD) is a leading cause of death and disability globally. In the United States, ASCVD accounts for one in three deaths, causing approximately 800,000 deaths each year^1^. Statins, cholesterol-lowering medications, are a vital tool in the prevention and treatment of ASCVD^2^. However, adherence to statin therapy remains low, especially among women and racial and ethnic minorities, raising significant concerns for long-term health disparities given the increased risk in mortality associated with non-adherence^3^.

There are multiple factors contributing to statin non-adherence. Studies have consistently shown that women, older adults, and historically marginalized populations are less likely to adhere to statins long-term^4^. Social determinants of health such as poverty, insurance status, and education level are also associated with non-adherence, with Black women without insurance facing the greatest challenges^5^. Health system factors, including provider knowledge and biases, similarly play a role, as inadequate patient-provider interactions, limited education, and short consultations hinder adherence^6^. While such evidence shows the complexity of factors that can influence statin adherence, the lack of detailed relevant social and systems data captured in electronic health records (EHRs) continue to limit our deeper understanding of key barriers, especially for underrepresented and vulnerable patient groups^7,8^.

The All of Us (AoU) dataset, a multi-domain database curated by the NIH’s Precision Medicine Initiative to address underrepresentation of racial and ethnic minorities, presents a unique opportunity to examine statin adherence across a diverse population. The objectives of this study are: first, to delineate disparities in statin adherence, and second, to gain valuable insights into the emergence of these disparities across diverse subpopulations. This understanding is essential for developing targeted interventions and policies that can address reasons underlying statin nonadherence and promote equitable cardiovascular health in diverse communities.

## METHODS

In this study, we perform a retrospective analysis to first assess how sociodemographic factors relate to statin adherence and second to characterize reasons for non-adherence utilizing the AoU cohort. This study was approved by the Institutional Review Board of Stanford University.

### Data source

We used the All of Us (AoU) v6 Controlled tier dataset, a multi-domain database from the NIH’s Precision Medicine Initiative^9^. The dataset integrates electronic health records (EHRs), physical measurements, biospecimens, and participants surveys. We analyzed three domains in the AoU dataset, the Demographic domain, The Drug Exposure domain, and the Healthcare Utilization Survey (HUS) domain.

### Study Population

The study cohort included individuals aged 18 or older prescribed a statin medication between 2017-2020 following the AoU v6 Controlled tier data cutoff (Overall Cohort). Statin prescriptions were identified using RxNorm codes, which included all generic and brand name statins (eTable1). To ensure that our study cohort and respective biometrics and survey responses represented participants accurately, we included only individuals with a statin prescription documentation within one year of the time of enrollment. Participants records lacking days supplied values or with suppressed information were excluded from the analysis. Participants were also excluded if their race was documented as “Asian” or “Other: American Indian/Alaskan Native, Middle Eastern, Asian, Native Hawaiian/ Pacific Islander” and sex at birth as “Other” due to the small sample sizes of these groups. We established a subset of individuals who responded to the health care utilization survey (HUS) to analyze adherence factors across racial and ethnic subgroups (Survey Cohort). We restricted participants to those who responded to the HUS question “Took Less Medicine to Save Money”, the survey with the highest response rate.

### Participants Characteristics

Participants’ data were captured at enrollment into the AoU cohort. The Demographic domain included date of birth, sex at birth, and self-reported race, ethnicity, and gender identities. Age was calculated as from the date of birth to time of enrollment. Race and ethnicity were categorized as Hispanic, Non-Hispanic White (NHW), and Non-Hispanic Black (NHB). Sex-at-birth was self-reported and included Male and Female. Other demographic variables including insurance status, socioeconomic status, educational attainment, and immigration status were obtained via self-reported demographic data.

Statin prescriptions was captured from the Drug Exposure domain, including fields on the number of days supplied, prescription start dates, and prescription names. Comorbidities such as myocardial infarction, liver disease, and diabetes were identified using ICD-9 and ICD-10 codes. The Charlson Comorbidity Index was calculated from EHRs. Mean values of lab measurements for high-density lipoprotein (HDL-C), low-density lipoprotein (LDL-C) cholesterol levels, and serum creatinine 12 months prior to the index date were included. Vital signs were obtained at the index date (See eTable 1 for data retrieval codes).

The Healthcare Utilization Survey (HUS) domain comprises qualitative responses about participants’ healthcare engagements. Questions evaluated the impact of access, utilization, and quality on individuals’ healthcare experiences and subsequent outcomes.^10^ Responses covered included 10 categories from the HUS, with thematically similar questions collapsed into survey variables (eTable 2). Experiencing any barrier within a category was coded as true, else it was coded as false. Nonresponses were assessed as false to account for missingness. K-means clustering was used for imputation of missing values.

### Outcome

The main outcome was percent days covered (PDC), calculated as the proportion of days in a 365-day period that an individual prescribed a statin obtained the prescription at the required dose. PDC is an established measure of adherence, approximating treatment access and use^11^. PDC was determined using the ‘days supplied’ value from prescription records. Adequate adherence was defined as PDC ≥ 0.80, based on established criteria from previous studies^47^. The time of study consent was used as the index date for each participant to align adherence metrics with recent participant data and survey responses.

### Statistical Analysis

The study was conducted in two phases. The first phase evaluated the association between demographics and statin adherence in overall cohort using Chi-square tests and Student’s t-tests. Univariate and multivariate regression models examined the association between sociodemographic factors and adequate adherence, adjusting for age, sex, and clinical characteristics.

The second phase involved examining reasons for statin adequate adherence compared to non-adherence (PDC<0.80%) in participants who completed the HUS. The cohort was stratified by racial and ethnic background to identify race and ethnic-specific reasons for non-adherence. Stepwise logistic regression modeling was used to identify predictors affecting statin adherence within each racial and ethnic subgroup. Sociodemographic and survey variables were included and the model was optimized for Akaike Information Criterion and model fit was assessed using the adjusted R-squared, with variable entry at p < 0.05 and removal at p > 0.10 to prevent overfitting. Models were compared across subgroups to identify differences in predictive profiles. All analyses were conducted on the AOURP platform using Python and R Workbench, with statistical significance defined as p < 0.05.

## RESULTS

### Overall Cohort

From a cohort of 372,380 participants, 69,529 individuals were prescribed statins (Figure 1). After applying inclusion and exclusion criteria —which excluded participants with missing or suppressed days supplied (46,119), those with no recorded usage within one year of consent (4,796), and study demographics with low sample sizes including other sex, other race, and Asian (1,585)— 17,029 participants with active statin prescriptions at the time of enrollment were included in the analysis. Of these, 7,786 completed the HUS (Survey Cohort). The mean age was 64.4 years (SD 11.5) and 50.2% were female at birth. Over two-thirds of the participants (67.1%) were Non-Hispanic White, 21.6% were Non-Hispanic Black, and 11.3% were Hispanic. Foreign-born participants accounted for 9.4% of the cohort (Table 1).

**Figure 1.**
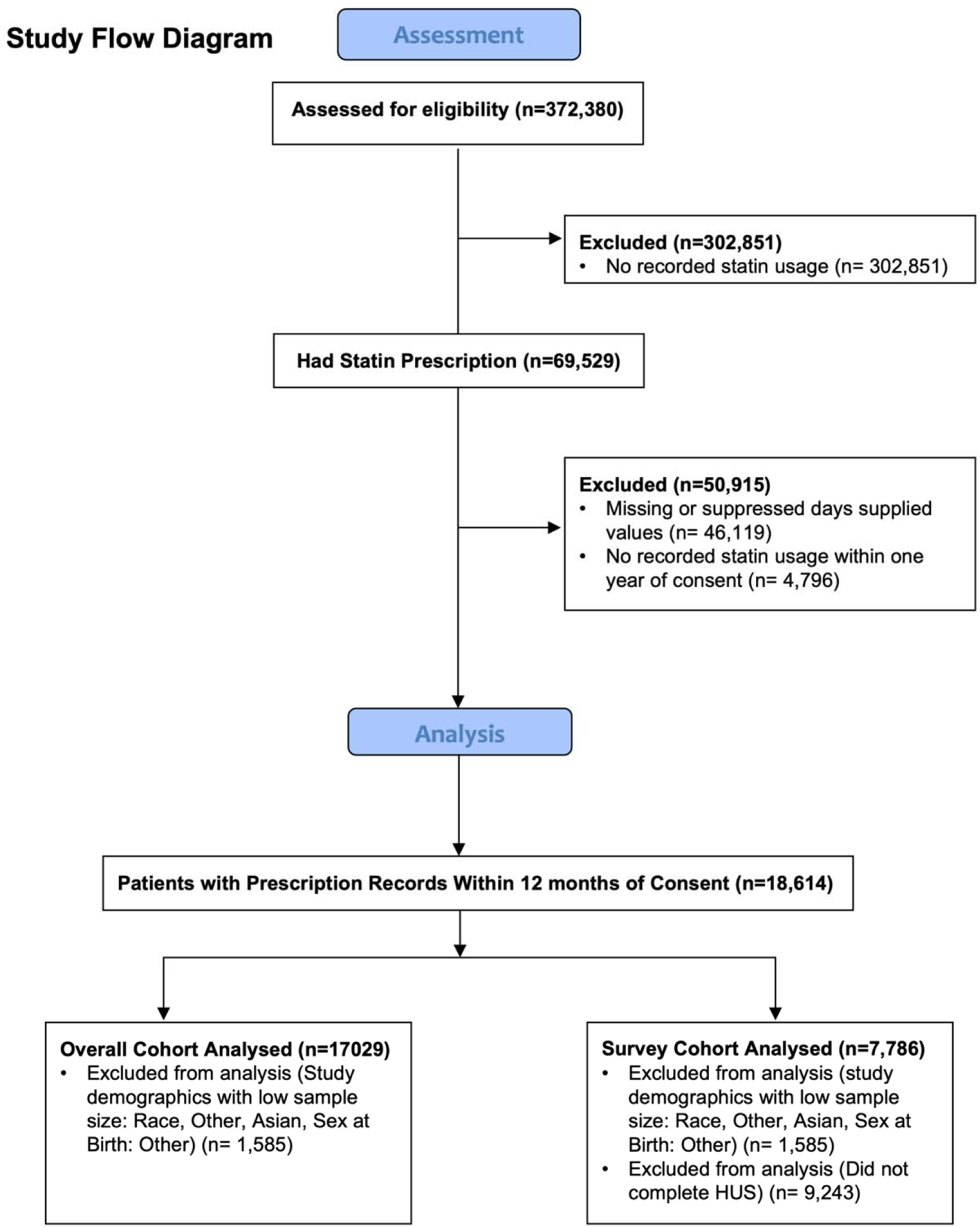
Study Consort Diagram. Exclusion criteria for the primary study population depended on differential data availability across study participants.

**Figure 2.**
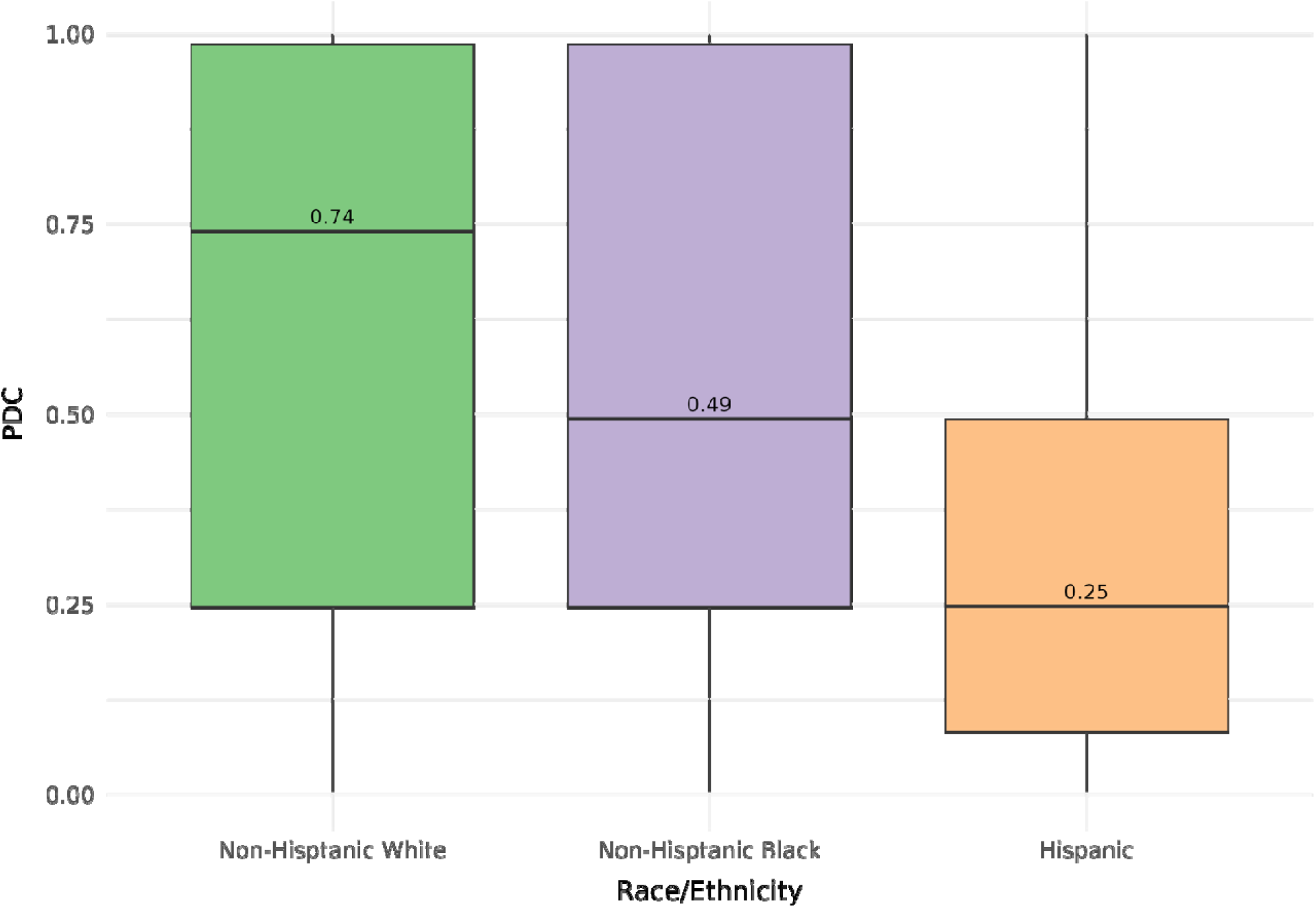
Distribution of PDC stratified by racial and ethnic subgroups. Stratified analysis revealed differences in median PDC (NHW: 0.740; NHB: 0.493; Hispanics: 0.247) when comparing racial and ethnic subgroups.

**Table 1.**
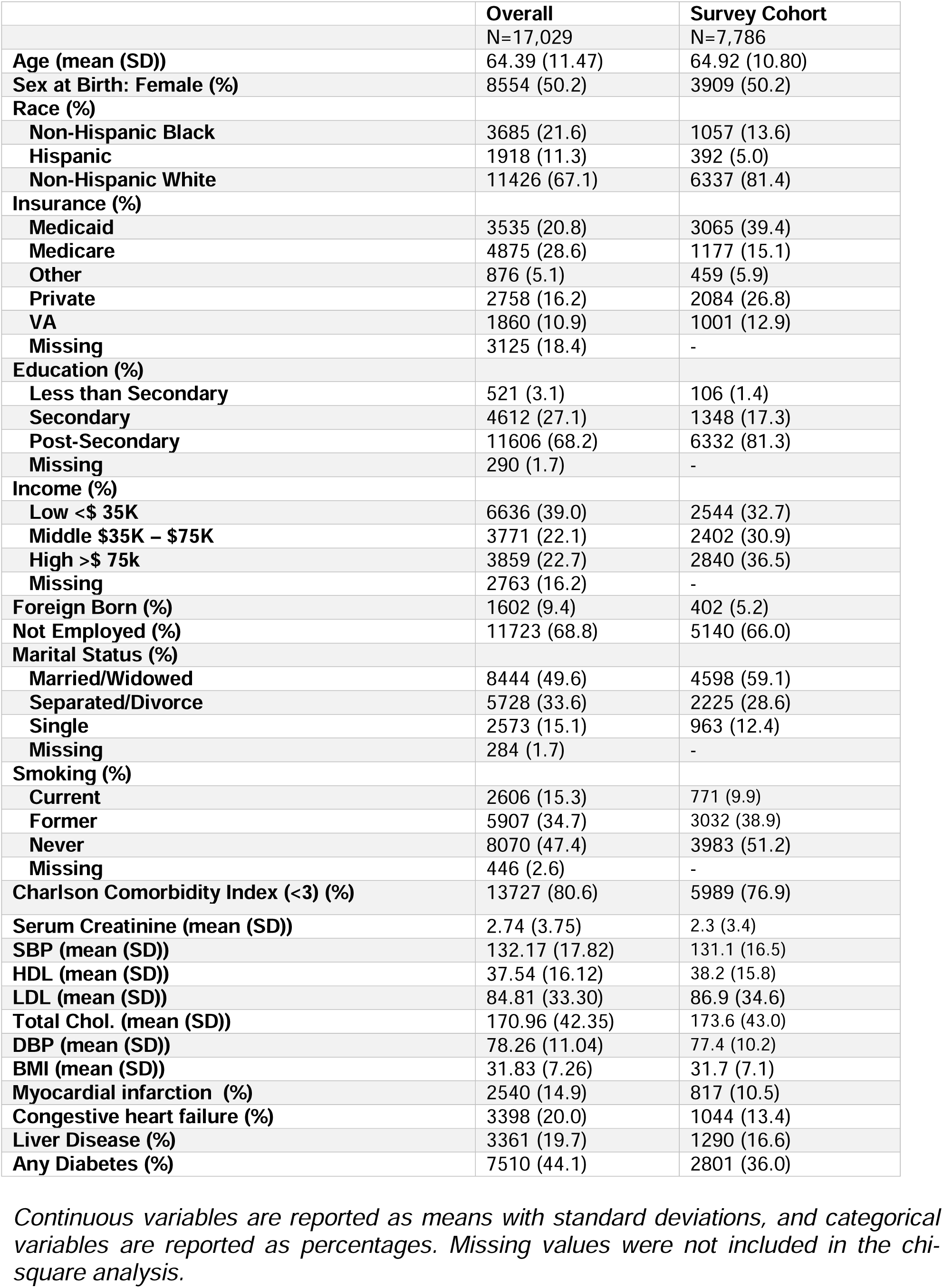
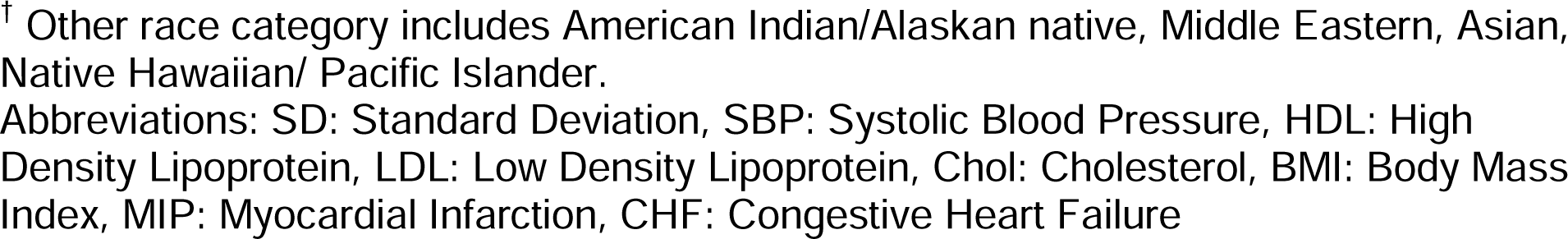
Baseline Characteristics of the Study Cohort.

**Table 2.**
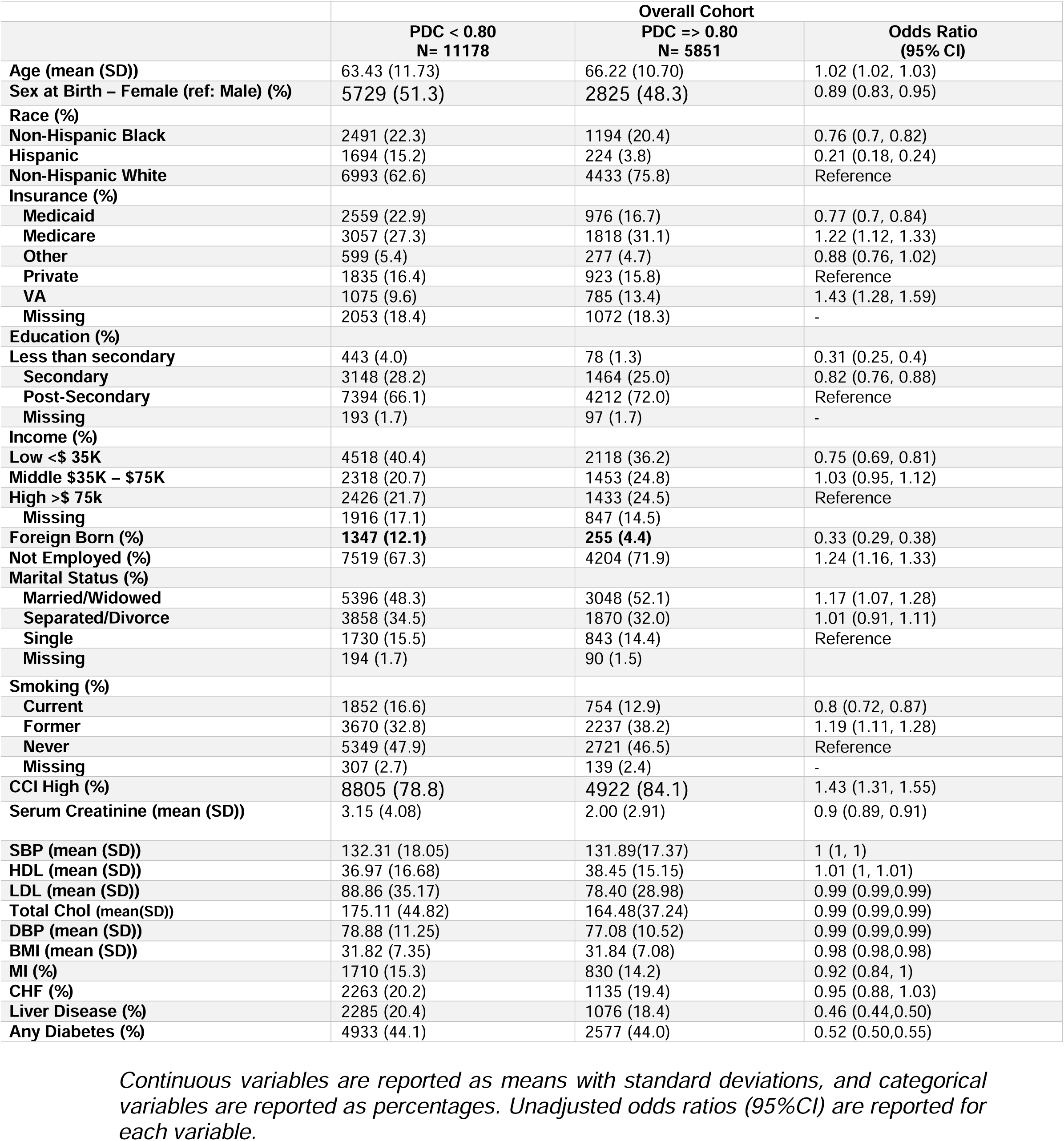
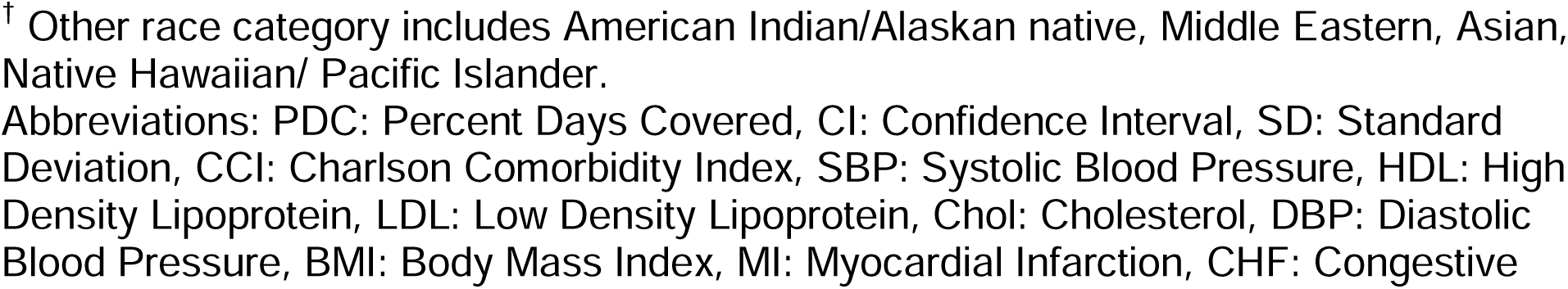
Bivariate Analysis of the Study Cohort According to Percent Days Covered (PDC) of Statin Prescription.

### Statin prescriptions

Among the 17,029 participants, the mean statin PDC was 57%, with 66% reporting a PDC ≤ 80%. Non-adherent participants were younger (63.4 ± 11.7 years vs. 66.2 ± 10.7 years), more female (51.3% vs 48.3%) had lower comorbidity burden (78.8% vs 84.1% with high comorbidities) and included a higher proportion of foreign-born individuals (12.1% vs 4.4%) compared to adherent participants as well as NHB (22.3% vs 20.4%) or Hispanic (15.2% vs. 3.8%) participants than adherent participants. When the cohort was stratified by racial and ethnic groups, NHW participants had a median PDC of 74% (IQR: [0.25,0.98]) Black participants had a PDC of 49% (IQR: [0.25,0.98]) and Hispanic participants had a PDC of 25% (IQR: [0.08,0.49]).

### Overall Cohort Analysis

The multivariable regression model identified factors associated with adequate statin adherence (Figure 3). Higher adherence was associated with being non-employed, having diabetes, or a Charlson index of 3 or higher (p>0.05). Lower adherence was observed among NHB (OR 0.75 (95% CI [0.68, 0.82]), and Hispanics (0.32 (95% CI [0.27, 0.38]) and immigrants (OR= 0.61, 95% CI: 0.51 to 0.72) compared to NHWs

**Figure 3.**
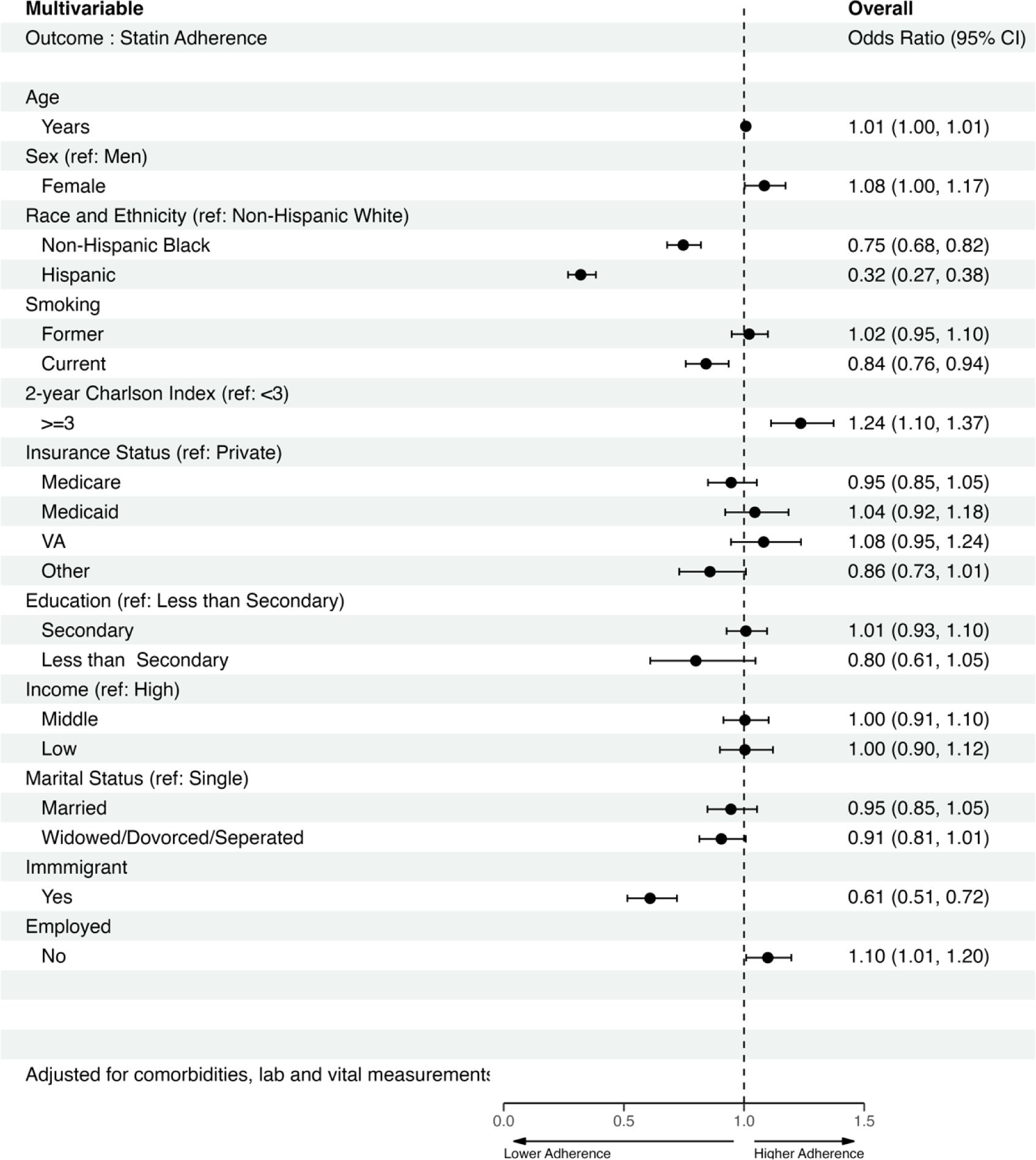
Multivariate Logistic Regression Analysis. *to Evaluate Factors Associated with Statin Adherence* in *O*verall *C*ohort Primary analysis included all baseline sociodemographic variables and was adjusted for clinical variables including SBP, DBP, LDL.

### Survey Cohort Analysis

In the sub-cohort of 7,786 participants who completed the HUS survey, the mean statin PDC was 0.64, and 60.8% had low adherence. The mean age of this sub cohort was 64.9 years (SD 10.80), and 81.4% were Non-Hispanic White, 13.6% Non-Hispanic Black, and 5.0% were Hispanic. Immigrant participants constituted 5.2% of the cohort. Survey participants were predominantly NHW, had fewer Medicare insured individuals, and exhibited higher education and income levels compared to the overall cohort (Table 1). Non-adherent survey participants shared characteristics with the overall non-adherent cohort, including more female, younger, Hispanic or NHB, lower income and education levels, immigrant status, and non-employed (eTable 3).

Multivariable analyses identified barriers to statin adherence unique to each race and ethnic group race-stratified (Table 3). Among NHW participants, employment barriers (OR 0.63, 95% CI [0.46, 0.86]) and provider inaccessibility (OR 0.56, 95% CI [0.40, 0.76]) were significantly associated with lower adherence. Higher adherence was associated with low (OR 1.18 CI 95% [1.03, 1.35]) and middle-income status (OR 1.17 CI 95% [1.04, 1.33]) compared to high income status. For NHB participants, patient anxiety (OR 0.53, 95% CI [0.30, 0.90]) and financial barriers (OR 0.65, 95% CI [0.50, 0.85]) were associated with lower adherence. Higher adherence (OR 2.38, 95% CI [1.17, 4.86]) was associated with less than a secondary education compared to post-secondary education. Among Hispanic participants, Patient anxiety (OR 0.14, 95% CI [0.02, 0.60]) and immigrant status (OR 0.36, 95% CI [0.17, 0.76]) were associated with lower adherence. Medicaid coverage compared to private insurance also predicted non-adherence (OR 0.26, 95% CI [0.11, 0.64]).

**Table 3:** Multivariable Stepwise Logistic Regression to Assess the Barriers For Non-Adherence Based on Survey Results.

| Multivariable Stepwise Logistic Regression Outcome: PDC >= 0.80 | OR (95% CI) | OR (95% CI) | OR (95% CI) |
| --- | --- | --- | --- |
|  | Non-Hispanic White | Non-Hispanic Black | Hispanic |
| **Survey Variables** | - | - | - |
| Employment Barrier* | **0.63 (0.46, 0.86)** |  |  |
| Provider Inaccessibility* | **0.56 (0.4, 0.76)** |  |  |
| Rural Barrier | 0.72 (0.48, 1.06) |  |  |
| Mutual Provider Relationship | 0.91 (0.81, 1.01) |  |  |
| Barriers Accessing Diverse HCW | 0.9 (0.79, 1.01) |  |  |
| Financial Barriers | 0.9 (0.81, 1) | **0.65 (0.5, 0.85)** |  |
| Patient Anxiety* |  | **0.53 (0.3, 0.9)** | **0.14 (0.02, 0.6)** |
| **Social Demographic Variables** |  |  |  |
| Insurance (ref: Private) |  |  |  |
| Medicare |  | 0.76 (0.51, 1.14) | 0.33 (0.1, 1.02) |
| Medicaid |  | 1.23 (0.87, 1.75) | **0.26 (0.11, 0.64)** |
| VA |  | 0.75 (0.4, 1.39) | **0.62 (0.2, 1.89)** |
| Other |  | 0.6 (0.3, 1.16) | 1.9 (0.6, 5.95) |
| Income (ref: High Income) |  |  |  |
| Low Income | 1.18 (1.03, 1.35) |  |  |
| Middle Income | 1.17 (1.04, 1.33) |  |  |
| Education (ref: Post-Secondary) |  |  |  |
| Less than Secondary |  | **2.46 (1.2, 5.17)** |  |
| Secondary |  | **1.33 (1, 1.76)** |  |
| Immigrant |  |  | **0.36 (0.17, 0.76)** |

*The input in the stepwise logistic regression model included all sociodemographic factors and comorbidities in addition to survey responses in each racial and ethnic subgroup. The stepwise model is adjusted for age and sex across all strata. Odds Ratios less than 1 indicate less likely to be adherent, Odds Ratios more than 1 indicate more likely to be adherent. \* Indicate statistically significant (p< 0.05) results.*
† Other race category includes American Indian/Alaskan Native, Middle Eastern, Asian, Native Hawaiian/ Pacific Islander.
Abbreviations: VA: Veterans Affairs, PDC: Percent Days Covered, CI: Confidence Interval

## Discussion

This national study using the AoU research cohort revealed low adherence to statin medications, with over two-thirds of the participants reporting adherence rates of 80% or less. Adherence was lower among racial and ethnic minorities. Key factors associated with adherence included employment status, comorbidity index, and immigration status, with the lowest adherence rates among NHB, Hispanic, and immigrant participants. We identified specific clusters of SDOH factors that vary significantly across racial and ethnic groups, playing a key role in adherence levels. This finding underscores complex and nuanced interactions between social and structural factors that affect health outcomes.

In examining statin adherence across different racial and ethnic groups within the AoU cohort, certain commonalities emerged. Age and gender consistently played a role in medication adherence, with elder and male participants demonstrating better adherence, regardless of racial or ethnic background, similar to other studies.^12^ Additionally, socioeconomic status, measured by insurance type and income level, was a shared determinant across all groups, a finding emphasized in separate studies advocating for the reduction of costs through insurance coverage or regulatory measures to improve medication adherence^4,13,14^. Patients with government-sponsored healthcare plans (e.g., Medicare or Veterans Health) had higher adherence rates than those with no insurance, suggesting insurance type may affect medication accessibility and affordability^15^.

For NHW participants, distinct factors influenced statin adherence that differed from other racial and ethnic minorities. Employment barriers, provider inaccessibility and rural living were notable issues, reflecting the influence of work-life demands and healthcare system navigation on medication management^16–18^. These barriers indicate that healthcare systems may be ill-equipped to support long-term and consistent care (e.g. medical advice or following-up appointments)^19,20^. Low healthcare workforce density in rural areas may exacerbate treatment access issues, hindering long-term care for chronic diseases like ASCVD ^17,21^. Improving adherence among NHW individuals may require prioritizing solutions to balance work-life demands with healthcare access and addressing workforce shortages. This could involve exploring telehealth services and implementing more flexible healthcare service hours.

Among NHB participants, patient anxiety emerged as a significant barrier to statin adherence, indicating that navigating complex health systems may discourage continued treatment engagement. Furthermore, NHB participants with less than secondary education showed higher statin adherence rates compared to NHW, possibly due to unmeasured factors, such as community support, cultural attitudes towards medication, and the dynamics of patient-physician communication^16,22,23^. Community-specific stressors also mediate negative health outcomes^24,25^. The Meyer Minority Stress model suggests higher adherence when patients trust their provider^26^. Trust in healthcare providers is crucial, as it influences patients’ willingness to follow medical advice and maintain a consistent treatment regime. Financial barriers were significant among NHB participants, which were associated with approximately 36% lower odds of adherence compared to those without financial barriers. Recent studies reveal that one in eight ASCVD patients, including Black Americans, face cost-related barriers to statins across, which disproportionately affect Black Americans^27–29^. This suggests that financial inaccessibility, symptomatic of broader social inequities, impacts Black participants’ access to and continuity of crucial preventative treatment.

For the Hispanic population within the AoU cohort, several factors impacted statin adherence. Immigration status was a significant predictor of adherence, with immigrants displaying notably lower adherence rates. Health literacy poses a major challenge for the broader U.S. population but is particularly pronounced among immigrant communities who face complex healthcare systems, language barriers and cultural differences^30–32^. Social safety insurance was also associated with lower adherence in this population, highlighting access gaps due to restricted eligibility requirements and complex case management. Lower adherence rates among immigrants may be partially due to self-selection effect, where healthier and more optimistic individuals perceive non-life-threatening issues as less urgent^33^. This is an important aspect to consider when comparing outcomes between other racial/ethnic groups.

Striking similarities between patient anxiety and lower adherence for both NHB and Hispanic participants underscore the role of trust in healthcare providers. Literature shows that a patient’s trust in their physician significantly influences their adherence to preventive measures.^23,34^ Distrust is common for both Hispanic and NHB individuals due to historic discrimination and racism in healthcare^35^. Public health innovations that strengthen bilingual communication, community-based support programs, and culturally competent healthcare providers can promote trust and confidence,^36^ enhancing adherence by addressing obstacles of continued treatment.

While this study provides novel insights, there are limitations. The AoU collects records from diverse data sources, leading to varying degrees of completeness and the potential for residual confounding. The observational nature of the analysis precludes causal inferences. In addition, the sample sizes of Asian and other racial/ethnic subgroups were limited, and only a subset completed the HUS entirely, necessitating exclusions for proper data analysis. Comparison of the survey cohort to the original cohort revealed notable differences in key indicators, including race, income, and education. This exclusion likely skews our results since the experience of social disadvantage and healthcare system barriers are closely tied to these socioeconomic characteristics. Regardless, the study’s strength lies in the utilization of the AoU cohort’s large and diverse participant cohort, which enhances the generalizability of the findings. The study population was racially, ethnically, and socioeconomically diverse - a level of diversity that has not been achieved by other studies on statin usage given limitations in EHR access.

### Public Health and clinical implications

It is becoming increasingly evident that the exclusive emphasis on race as a determinant of health disparities restricts our ability to identify and address the modifiable structural barriers that underlie them^36^. These findings emphasize such inadequacy, highlighting the varied effects of the SDoH across different patient populations, underscoring the ongoing need to change how race and ethnicity are discussed and addressed in medicine. Tackling medication nonadherence requires recognizing social and economic inequities that impact patients’ lives. Ensuring consistent, high quality healthcare access for everyone, regardless of background, is crucial, especially given the potential of preventive measures like statins to reduce disparities and cardiovascular disease incidence. Refining how systems interact with diverse communities can cultivate better care touchpoints. Address historical distrust in healthcare systems could also engage in broader societal discussions about dismantling systemic injustices. Recognize that health disparities are deeply intertwined with broader social, economic, and political factors calls for a holistic approach.

In conclusion, this national study has illuminated the intricate mosaic of factors influencing statin adherence across a racially and ethnically diverse cohort, highlighting the need for multifaceted and culturally sensitive interventions. While common factors such as age, gender, and socioeconomic status universally affect statin adherence each group’s challenges are uniquely shaped by their experiences and social determinants. To improve statin adherence across all populations, interventions must address shared challenges and consider diverse social, cultural, and economic realities. Future research should continue to investigate how social conditions and experiences intersect with medication adherence, paving the way for policy interventions tailored to these needs. As we move towards a more inclusive healthcare model, the insights from this study provide a valuable roadmap for developing precise, targeted strategies that cater to the nuanced needs of every patient population.

## Data Availability

The data utilized in this study are available through the All of Us Research Program website. Researchers can access the dataset by visiting All of Us and following the appropriate procedures for data access and use. All relevant data supporting the findings of this study are included in the article or can be obtained from the All of Us platform.

https://allofus.nih.gov/

## Acknowledgements

We thank the All of Us participants for their participation in this study.

## Declaration of interests

Dr. Hernandez-Boussard had full access to all of the data in the study and takes responsibility for the integrity of the data and the accuracy of the data analysis. She attests that all listed authors meet authorship criteria and that no others meeting the criteria have been omitted. She further affirms that the manuscript is an honest, accurate, and transparent account of the study being reported; that no important aspects of the study have been omitted; and that any discrepancies from the study as planned have been explained.

## Contributors

Study concept and design: THB, GE, ZA, AdH

Acquisition of data: THB, GE, ZA, AL

Analysis and interpretation of data: THB, GE, ZA, AdH, AL, MN, FR

Drafting of the manuscript: GE, AdH, ZA

Critical revision of the manuscript for important intellectual content: all authors.

Statistical analysis: GE, ZA, AdH

Administrative, technical or material support: THB.

Study supervision: THB.

## Funding

Dr Hernandez-Boussard was supported, in part, by the National Library Of Medicine of the National Institutes of Health under Award Number R01LM013362. Dr. Rodriguez was funded by grants from the NIH National Heart, Lung, and Blood Institute (1K01HL144607; R01HL168188) and the Doris Duke Foundation (Grant #2022051). The content is solely the responsibility of the authors and does not necessarily represent the official views of the National Institutes of Health.

## Ethical approval

The study was approved by the local ethics committee.

## Disclosures

Dr. Hernandez-Boussard reports consulting fees from Grai-Matter, Paul Hartmann AG, outside the submitted work and she is a board member of Athelo Health

Dr. Rodriguez reports consulting fees from HealthPals, Novartis, NovoNordisk, Esperion Therapeutics, Movano Health, Kento Health, Inclusive Health, Edwards, Arrowhead Pharmaceuticals, HeartFlow, and iRhythm outside the submitted work.

## Notes

### Competing Interest Statement

The authors have declared no competing interest.

### Author Declarations

This study presents aggregate data and was approved by the Institutional Review Board of Stanford University.

