## Supplement Material for "Racial and Ethnic Disparities in Statin Adherence: Insights from the All of Us Research Program"

**Supplementary Online Content**

eTable 1. Associated codes utilized in data queries in the AoURP

eTable 2. Health Utilization Survey Question Categorizations

eTable 3: Bivariate Analysis of the Survey Cohort According to Percent Days Covered (PDC) of Statin Prescription

**eTable 1. Associated codes utilized in data queries in the AoURP**

| **Prescription Name** | **Type** | **Query Code** |
| --- | --- | --- |
| Lovastatin | RxNorm | 6472 |
| Pitavastatin | RxNorm | 861634 |
| Fluvastatin | RxNorm | 41127 |
| Rosuvastatin | RxNorm | 301542 |
| Pravastatin | RxNorm | 42463 |
| Atorvastatin | RxNorm | 83367 |
| Simvastatinn | RxNorm | 36567 |
| **Regression Variables** | **Type** | **Query Code** |
| Age | AoU Source | date_of_birh |
| Smoking Status | AoU Source | 1586182 |
| Educational Attainment | AoU Source | 1585940 |
| Income | AoU Source | 1585375 |
| Sex at Birth | AoU Source | 1585845 |
| Insurance | AoU Source | 1585386 |
| Birthplace | AoU Source | 1586135 |
| Race/ethnicity | AoU Source | 1586140 |
| Employment Status | AoU Source | 1585952 |
| Marital Status | AoU Source | 1585892 |
| Healthcare Utilization Survey | AoU Source | 43528895 |

| **Disease Name** | **Type** | **Code** | **Disease Name** | **Type** | **Code** |
| --- | --- | --- | --- | --- | --- |
| Myocardial Infarction | ICD9CM | 410 | Mild Liver Disease | ICD10 | B18 |
| Myocardial Infarction | ICD9CM | 412 | Mild Liver Disease | ICD10 | K73 |
| Congestive Heart Failure | ICD9CM | 398.91 | Mild Liver Disease | ICD10 | K74 |
| Congestive Heart Failure | ICD9CM | 402.01 | Mild Liver Disease | ICD10 | K70.0 |
| Congestive Heart Failure | ICD9CM | 402.11 | Mild Liver Disease | ICD10 | K70.1 |
| Congestive Heart Failure | ICD9CM | 402.91 | Mild Liver Disease | ICD10 | K70.2 |
| Congestive Heart Failure | ICD9CM | 404.01 | Mild Liver Disease | ICD10 | K70.3 |
| Congestive Heart Failure | ICD9CM | 404.03 | Mild Liver Disease | ICD10 | K70.9 |
| Congestive Heart Failure | ICD9CM | 404.11 | Mild Liver Disease | ICD10 | K71.7 |
| Congestive Heart Failure | ICD9CM | 404.13 | Mild Liver Disease | ICD10 | K71.3 |
| Congestive Heart Failure | ICD9CM | 404.91 | Mild Liver Disease | ICD10 | K71.4 |
| Congestive Heart Failure | ICD9CM | 404.93 | Mild Liver Disease | ICD10 | K71.5 |
| Congestive Heart Failure | ICD9CM | 425.4 | Mild Liver Disease | ICD10 | K76.0 |
| Congestive Heart Failure | ICD9CM | 425.5 | Mild Liver Disease | ICD10 | K76.2 |
| Congestive Heart Failure | ICD9CM | 425.7 | Mild Liver Disease | ICD10 | K76.3 |
| Congestive Heart Failure | ICD9CM | 425.8 | Mild Liver Disease | ICD10 | K76.4 |
| Congestive Heart Failure | ICD9CM | 425.9 | Mild Liver Disease | ICD10 | K76.8 |
| Congestive Heart Failure | ICD9CM | 428 | Mild Liver Disease | ICD10 | K76.9 |
| Periphral Vascular Disease | ICD9CM | 930 | Mild Liver Disease | ICD10 | Z94.4 |
| Periphral Vascular Disease | ICD9CM | 437.3 | Diabetes without complications | ICD10 | E10.0 |
| Periphral Vascular Disease | ICD9CM | 440 | Diabetes without complications | ICD10 | E10.1 |
| Periphral Vascular Disease | ICD9CM | 441 | Diabetes without complications | ICD10 | E10.6 |
| Periphral Vascular Disease | ICD9CM | 443.1 | Diabetes without complications | ICD10 | E10.8 |
| Periphral Vascular Disease | ICD9CM | 443.2 | Diabetes without complications | ICD10 | E10.9 |
| Periphral Vascular Disease | ICD9CM | 443.8 | Diabetes without complications | ICD10 | E11.0 |
| Periphral Vascular Disease | ICD9CM | 443.9 | Diabetes without complications | ICD10 | E11.1 |
| Periphral Vascular Disease | ICD9CM | 447.1 | Diabetes without complications | ICD10 | E11.6 |
| Periphral Vascular Disease | ICD9CM | 557.1 | Diabetes without complications | ICD10 | E11.8 |
| Periphral Vascular Disease | ICD9CM | 557.9 | Diabetes without complications | ICD10 | E11.9 |
| Periphral Vascular Disease | ICD9CM | V43.4 | Diabetes without complications | ICD10 | E12.0 |
| Cerebrovascular Disease | ICD9CM | 362.34 | Diabetes without complications | ICD10 | E12.1 |
| Cerebrovascular Disease | ICD9CM | 430 | Diabetes without complications | ICD10 | E12.6 |
| Cerebrovascular Disease | ICD9CM | 431 | Diabetes without complications | ICD10 | E12.8 |
| Cerebrovascular Disease | ICD9CM | 432 | Diabetes without complications | ICD10 | E12.9 |
| Cerebrovascular Disease | ICD9CM | 433 | Diabetes without complications | ICD10 | E13.0 |
| Cerebrovascular Disease | ICD9CM | 434 | Diabetes without complications | ICD10 | E13.1 |
| Cerebrovascular Disease | ICD9CM | 435 | Diabetes without complications | ICD10 | E13.6 |
| Cerebrovascular Disease | ICD9CM | 436 | Diabetes without complications | ICD10 | E13.8 |
| Cerebrovascular Disease | ICD9CM | 437 | Diabetes without complications | ICD10 | E13.9 |
| Cerebrovascular Disease | ICD9CM | 438 | Diabetes without complications | ICD10 | E14.0 |
| Dementia | ICD9CM | 290 | Diabetes without complications | ICD10 | E14.1 |
| Dementia | ICD9CM | 294.1 | Diabetes without complications | ICD10 | E14.6 |
| Dementia | ICD9CM | 331.2 | Diabetes without complications | ICD10 | E14.8 |
| Chronic Pulmonary Disease | ICD9CM | 416.8 | Diabetes without complications | ICD10 | E14.9 |
| Chronic Pulmonary Disease | ICD9CM | 416.9 | Diabetes with complications | ICD10 | E10.2 |
| Chronic Pulmonary Disease | ICD9CM | 490 | Diabetes with complications | ICD10 | E10.3 |
| Chronic Pulmonary Disease | ICD9CM | 491 | Diabetes with complications | ICD10 | E10.4 |
| Chronic Pulmonary Disease | ICD9CM | 492 | Diabetes with complications | ICD10 | E10.5 |
| Chronic Pulmonary Disease | ICD9CM | 493 | Diabetes with complications | ICD10 | E10.7 |
| Chronic Pulmonary Disease | ICD9CM | 494 | Diabetes with complications | ICD10 | E11.2 |
| Chronic Pulmonary Disease | ICD9CM | 495 | Diabetes with complications | ICD10 | E11.3 |
| Chronic Pulmonary Disease | ICD9CM | 496 | Diabetes with complications | ICD10 | E11.4 |
| Chronic Pulmonary Disease | ICD9CM | 500 | Diabetes with complications | ICD10 | E11.5 |
| Chronic Pulmonary Disease | ICD9CM | 501 | Diabetes with complications | ICD10 | E11.7 |
| Chronic Pulmonary Disease | ICD9CM | 502 | Diabetes with complications | ICD10 | E12.2 |
| Chronic Pulmonary Disease | ICD9CM | 503 | Diabetes with complications | ICD10 | E12.3 |
| Chronic Pulmonary Disease | ICD9CM | 504 | Diabetes with complications | ICD10 | E12.4 |
| Chronic Pulmonary Disease | ICD9CM | 505 | Diabetes with complications | ICD10 | E12.5 |
| Chronic Pulmonary Disease | ICD9CM | 506.4 | Diabetes with complications | ICD10 | E12.7 |
| Chronic Pulmonary Disease | ICD9CM | 508.1 | Diabetes with complications | ICD10 | E13.2 |
| Chronic Pulmonary Disease | ICD9CM | 508.8 | Diabetes with complications | ICD10 | E13.3 |
| Connective Tissue Disease-Rheumatic Disease | ICD9CM | 446.5 | Diabetes with complications | ICD10 | E13.4 |
| Connective Tissue Disease-Rheumatic Disease | ICD9CM | 710 | Diabetes with complications | ICD10 | E13.5 |
| Connective Tissue Disease-Rheumatic Disease | ICD9CM | 710.1 | Diabetes with complications | ICD10 | E13.7 |
| Connective Tissue Disease-Rheumatic Disease | ICD9CM | 710.2 | Diabetes with complications | ICD10 | E14.2 |
| Connective Tissue Disease-Rheumatic Disease | ICD9CM | 710.3 | Diabetes with complications | ICD10 | E14.3 |
| Connective Tissue Disease-Rheumatic Disease | ICD9CM | 710.4 | Diabetes with complications | ICD10 | E14.4 |
| Connective Tissue Disease-Rheumatic Disease | ICD9CM | 714 | Diabetes with complications | ICD10 | E14.5 |
| Connective Tissue Disease-Rheumatic Disease | ICD9CM | 714.1 | Diabetes with complications | ICD10 | E14.7 |
| Connective Tissue Disease-Rheumatic Disease | ICD9CM | 714.2 | Paraplegia and Hemiplegia | ICD10 | G81 |
| Connective Tissue Disease-Rheumatic Disease | ICD9CM | 714.8 | Paraplegia and Hemiplegia | ICD10 | G82 |
| Connective Tissue Disease-Rheumatic Disease | ICD9CM | 725 | Paraplegia and Hemiplegia | ICD10 | G04.1 |
| Peptic Ulcer Disease | ICD9CM | 531 | Paraplegia and Hemiplegia | ICD10 | G11.4 |
| Peptic Ulcer Disease | ICD9CM | 532 | Paraplegia and Hemiplegia | ICD10 | G80.1 |
| Peptic Ulcer Disease | ICD9CM | 533 | Paraplegia and Hemiplegia | ICD10 | G80.2 |
| Peptic Ulcer Disease | ICD9CM | 534 | Paraplegia and Hemiplegia | ICD10 | G83.0 |
| Mild Liver Disease | ICD9CM | 702.2 | Paraplegia and Hemiplegia | ICD10 | G83.1 |
| Mild Liver Disease | ICD9CM | 702.3 | Paraplegia and Hemiplegia | ICD10 | G83.2 |
| Mild Liver Disease | ICD9CM | 703.2 | Paraplegia and Hemiplegia | ICD10 | G83.3 |
| Mild Liver Disease | ICD9CM | 703.3 | Paraplegia and Hemiplegia | ICD10 | G83.4 |
| Mild Liver Disease | ICD9CM | 704.4 | Paraplegia and Hemiplegia | ICD10 | G83.9 |
| Mild Liver Disease | ICD9CM | 705.4 | Renal Disease | ICD10 | N18 |
| Mild Liver Disease | ICD9CM | 706 | Renal Disease | ICD10 | N19 |
| Mild Liver Disease | ICD9CM | 709 | Renal Disease | ICD10 | N05.2 |
| Mild Liver Disease | ICD9CM | 570 | Renal Disease | ICD10 | N05.3 |
| Mild Liver Disease | ICD9CM | 571 | Renal Disease | ICD10 | N05.4 |
| Mild Liver Disease | ICD9CM | 573.3 | Renal Disease | ICD10 | N05.5 |
| Mild Liver Disease | ICD9CM | 573.4 | Renal Disease | ICD10 | N05.6 |
| Mild Liver Disease | ICD9CM | 573.8 | Renal Disease | ICD10 | N05.7 |
| Mild Liver Disease | ICD9CM | 573.9 | Renal Disease | ICD10 | N25.0 |
| Mild Liver Disease | ICD9CM | V42.7 | Renal Disease | ICD10 | I12.0 |
| Diabetes without complications | ICD9CM | 250 | Renal Disease | ICD10 | I13.1 |
| Diabetes without complications | ICD9CM | 250.1 | Renal Disease | ICD10 | N03.2 |
| Diabetes without complications | ICD9CM | 250.2 | Renal Disease | ICD10 | N03.3 |
| Diabetes without complications | ICD9CM | 250.3 | Renal Disease | ICD10 | N03.4 |
| Diabetes without complications | ICD9CM | 250.8 | Renal Disease | ICD10 | N03.5 |
| Diabetes without complications | ICD9CM | 250.9 | Renal Disease | ICD10 | N03.6 |
| Diabetes with complications | ICD9CM | 250.4 | Renal Disease | ICD10 | N03.7 |
| Diabetes with complications | ICD9CM | 250.5 | Renal Disease | ICD10 | Z49.0 |
| Diabetes with complications | ICD9CM | 250.6 | Renal Disease | ICD10 | Z49.1 |
| Diabetes with complications | ICD9CM | 250.7 | Renal Disease | ICD10 | Z49.2 |
| Paraplegia and Hemiplegia | ICD9CM | 334.1 | Renal Disease | ICD10 | Z94.0 |
| Paraplegia and Hemiplegia | ICD9CM | 342 | Renal Disease | ICD10 | Z99.2 |
| Paraplegia and Hemiplegia | ICD9CM | 343 | Cancer | ICD10 | C00 |
| Paraplegia and Hemiplegia | ICD9CM | 344 | Cancer | ICD10 | C01 |
| Paraplegia and Hemiplegia | ICD9CM | 344.1 | Cancer | ICD10 | C02 |
| Paraplegia and Hemiplegia | ICD9CM | 344.2 | Cancer | ICD10 | C03 |
| Paraplegia and Hemiplegia | ICD9CM | 344.3 | Cancer | ICD10 | C04 |
| Paraplegia and Hemiplegia | ICD9CM | 344.4 | Cancer | ICD10 | C05 |
| Paraplegia and Hemiplegia | ICD9CM | 344.5 | Cancer | ICD10 | C06 |
| Paraplegia and Hemiplegia | ICD9CM | 344.6 | Cancer | ICD10 | C07 |
| Paraplegia and Hemiplegia | ICD9CM | 344.9 | Cancer | ICD10 | C08 |
| Renal Disease | ICD9CM | 403.01 | Cancer | ICD10 | C09 |
| Renal Disease | ICD9CM | 403.11 | Cancer | ICD10 | C10 |
| Renal Disease | ICD9CM | 403.91 | Cancer | ICD10 | C11 |
| Renal Disease | ICD9CM | 404.02 | Cancer | ICD10 | C12 |
| Renal Disease | ICD9CM | 404.03 | Cancer | ICD10 | C13 |
| Renal Disease | ICD9CM | 404.12 | Cancer | ICD10 | C14 |
| Renal Disease | ICD9CM | 404.13 | Cancer | ICD10 | C15 |
| Renal Disease | ICD9CM | 404.92 | Cancer | ICD10 | C16 |
| Renal Disease | ICD9CM | 404.93 | Cancer | ICD10 | C17 |
| Renal Disease | ICD9CM | 582 | Cancer | ICD10 | C18 |
| Renal Disease | ICD9CM | 583 | Cancer | ICD10 | C19 |
| Renal Disease | ICD9CM | 583.1 | Cancer | ICD10 | C20 |
| Renal Disease | ICD9CM | 583.2 | Cancer | ICD10 | C21 |
| Renal Disease | ICD9CM | 583.4 | Cancer | ICD10 | C22 |
| Renal Disease | ICD9CM | 583.6 | Cancer | ICD10 | C23 |
| Renal Disease | ICD9CM | 583.7 | Cancer | ICD10 | C24 |
| Renal Disease | ICD9CM | 585 | Cancer | ICD10 | C25 |
| Renal Disease | ICD9CM | 586 | Cancer | ICD10 | C26 |
| Renal Disease | ICD9CM | 588 | Cancer | ICD10 | C30 |
| Renal Disease | ICD9CM | V42.0 | Cancer | ICD10 | C31 |
| Renal Disease | ICD9CM | V45.1 | Cancer | ICD10 | C32 |
| Renal Disease | ICD9CM | V56 | Cancer | ICD10 | C33 |
| Cancer | ICD9CM | 140 | Cancer | ICD10 | C34 |
| Cancer | ICD9CM | 141 | Cancer | ICD10 | C37 |
| Cancer | ICD9CM | 142 | Cancer | ICD10 | C38 |
| Cancer | ICD9CM | 143 | Cancer | ICD10 | C39 |
| Cancer | ICD9CM | 144 | Cancer | ICD10 | C40 |
| Cancer | ICD9CM | 145 | Cancer | ICD10 | C41 |
| Cancer | ICD9CM | 146 | Cancer | ICD10 | C43 |
| Cancer | ICD9CM | 147 | Cancer | ICD10 | C45 |
| Cancer | ICD9CM | 148 | Cancer | ICD10 | C46 |
| Cancer | ICD9CM | 149 | Cancer | ICD10 | C47 |
| Cancer | ICD9CM | 150 | Cancer | ICD10 | C48 |
| Cancer | ICD9CM | 151 | Cancer | ICD10 | C49 |
| Cancer | ICD9CM | 152 | Cancer | ICD10 | C50 |
| Cancer | ICD9CM | 153 | Cancer | ICD10 | C51 |
| Cancer | ICD9CM | 154 | Cancer | ICD10 | C52 |
| Cancer | ICD9CM | 155 | Cancer | ICD10 | C53 |
| Cancer | ICD9CM | 156 | Cancer | ICD10 | C54 |
| Cancer | ICD9CM | 157 | Cancer | ICD10 | C55 |
| Cancer | ICD9CM | 158 | Cancer | ICD10 | C56 |
| Cancer | ICD9CM | 159 | Cancer | ICD10 | C57 |
| Cancer | ICD9CM | 160 | Cancer | ICD10 | C58 |
| Cancer | ICD9CM | 161 | Cancer | ICD10 | C60 |
| Cancer | ICD9CM | 162 | Cancer | ICD10 | C61 |
| Cancer | ICD9CM | 163 | Cancer | ICD10 | C62 |
| Cancer | ICD9CM | 164 | Cancer | ICD10 | C63 |
| Cancer | ICD9CM | 165 | Cancer | ICD10 | C64 |
| Cancer | ICD9CM | 170 | Cancer | ICD10 | C65 |
| Cancer | ICD9CM | 171 | Cancer | ICD10 | C66 |
| Cancer | ICD9CM | 172 | Cancer | ICD10 | C67 |
| Cancer | ICD9CM | 174 | Cancer | ICD10 | C68 |
| Cancer | ICD9CM | 175 | Cancer | ICD10 | C69 |
| Cancer | ICD9CM | 176 | Cancer | ICD10 | C70 |
| Cancer | ICD9CM | 179 | Cancer | ICD10 | C71 |
| Cancer | ICD9CM | 180 | Cancer | ICD10 | C72 |
| Cancer | ICD9CM | 181 | Cancer | ICD10 | C73 |
| Cancer | ICD9CM | 182 | Cancer | ICD10 | C74 |
| Cancer | ICD9CM | 183 | Cancer | ICD10 | C75 |
| Cancer | ICD9CM | 184 | Cancer | ICD10 | C76 |
| Cancer | ICD9CM | 185 | Cancer | ICD10 | C81 |
| Cancer | ICD9CM | 186 | Cancer | ICD10 | C82 |
| Cancer | ICD9CM | 187 | Cancer | ICD10 | C83 |
| Cancer | ICD9CM | 188 | Cancer | ICD10 | C84 |
| Cancer | ICD9CM | 189 | Cancer | ICD10 | C85 |
| Cancer | ICD9CM | 190 | Cancer | ICD10 | C88 |
| Cancer | ICD9CM | 191 | Cancer | ICD10 | C90 |
| Cancer | ICD9CM | 192 | Cancer | ICD10 | C91 |
| Cancer | ICD9CM | 193 | Cancer | ICD10 | C92 |
| Cancer | ICD9CM | 194 | Cancer | ICD10 | C93 |
| Cancer | ICD9CM | 195 | Cancer | ICD10 | C94 |
| Cancer | ICD9CM | 200 | Cancer | ICD10 | C95 |
| Cancer | ICD9CM | 201 | Cancer | ICD10 | C96 |
| Cancer | ICD9CM | 202 | Cancer | ICD10 | C97 |
| Cancer | ICD9CM | 203 | Moderate or Severe Liver Disease | ICD10 | K70.4 |
| Cancer | ICD9CM | 204 | Moderate or Severe Liver Disease | ICD10 | K71.1 |
| Cancer | ICD9CM | 205 | Moderate or Severe Liver Disease | ICD10 | K72.1 |
| Cancer | ICD9CM | 206 | Moderate or Severe Liver Disease | ICD10 | K72.9 |
| Cancer | ICD9CM | 207 | Moderate or Severe Liver Disease | ICD10 | K76.5 |
| Cancer | ICD9CM | 208 | Moderate or Severe Liver Disease | ICD10 | K76.6 |
| Cancer | ICD9CM | 238.6 | Moderate or Severe Liver Disease | ICD10 | K76.7 |
| Moderate or Severe Liver Disease | ICD9CM | 456 | Moderate or Severe Liver Disease | ICD10 | I85.0 |
| Moderate or Severe Liver Disease | ICD9CM | 456.1 | Moderate or Severe Liver Disease | ICD10 | I85.9 |
| Moderate or Severe Liver Disease | ICD9CM | 456.2 | Moderate or Severe Liver Disease | ICD10 | I86.4 |
| Moderate or Severe Liver Disease | ICD9CM | 572.2 | Moderate or Severe Liver Disease | ICD10 | I98.2 |
| Moderate or Severe Liver Disease | ICD9CM | 572.3 | Metastatic Carcinoma | ICD10 | C77 |
| Moderate or Severe Liver Disease | ICD9CM | 572.4 | Metastatic Carcinoma | ICD10 | C78 |
| Moderate or Severe Liver Disease | ICD9CM | 572.8 | Metastatic Carcinoma | ICD10 | C79 |
| Metastatic Carcinoma | ICD9CM | 196 | Metastatic Carcinoma | ICD10 | C80 |
| Metastatic Carcinoma | ICD9CM | 197 | AIDS/HIV | ICD10 | B20 |
| Metastatic Carcinoma | ICD9CM | 198 | AIDS/HIV | ICD10 | B21 |
| Metastatic Carcinoma | ICD9CM | 199 | AIDS/HIV | ICD10 | B22 |
| AIDS/HIV | ICD9CM | 42 | AIDS/HIV | ICD10 | B24 |
| AIDS/HIV | ICD9CM | 43 | Cerebrovascular Disease | ICD10 | G45 |
| AIDS/HIV | ICD9CM | 44 | Cerebrovascular Disease | ICD10 | G46 |
| Myocardial Infarction | ICD10 | I21 | Cerebrovascular Disease | ICD10 | I60 |
| Myocardial Infarction | ICD10 | I22 | Cerebrovascular Disease | ICD10 | I61 |
| Myocardial Infarction | ICD10 | I25.2 | Cerebrovascular Disease | ICD10 | I62 |
| Congestive Heart Failure | ICD10 | I43 | Cerebrovascular Disease | ICD10 | I63 |
| Congestive Heart Failure | ICD10 | I50 | Cerebrovascular Disease | ICD10 | I64 |
| Congestive Heart Failure | ICD10 | I09.9 | Cerebrovascular Disease | ICD10 | I65 |
| Congestive Heart Failure | ICD10 | I11.0 | Cerebrovascular Disease | ICD10 | I66 |
| Congestive Heart Failure | ICD10 | I13.0 | Cerebrovascular Disease | ICD10 | I67 |
| Congestive Heart Failure | ICD10 | I13.2 | Cerebrovascular Disease | ICD10 | I68 |
| Congestive Heart Failure | ICD10 | I25.5 | Cerebrovascular Disease | ICD10 | I69 |
| Congestive Heart Failure | ICD10 | I42.0 | Cerebrovascular Disease | ICD10 | H34.0 |
| Congestive Heart Failure | ICD10 | I42.5 | Dementia | ICD10 | F00 |
| Congestive Heart Failure | ICD10 | I42.6 | Dementia | ICD10 | F01 |
| Congestive Heart Failure | ICD10 | I42.7 | Dementia | ICD10 | F02 |
| Congestive Heart Failure | ICD10 | I42.8 | Dementia | ICD10 | F03 |
| Congestive Heart Failure | ICD10 | I42.9 | Dementia | ICD10 | G30 |
| Congestive Heart Failure | ICD10 | P29.0 | Dementia | ICD10 | F05.1 |
| Periphral Vascular Disease | ICD10 | I70 | Dementia | ICD10 | G31.1 |
| Periphral Vascular Disease | ICD10 | I71 | Chronic Pulmonary Disease | ICD10 | J40 |
| Periphral Vascular Disease | ICD10 | I73.1 | Chronic Pulmonary Disease | ICD10 | J41 |
| Periphral Vascular Disease | ICD10 | I73.8 | Chronic Pulmonary Disease | ICD10 | J42 |
| Periphral Vascular Disease | ICD10 | I73.9 | Chronic Pulmonary Disease | ICD10 | J43 |
| Periphral Vascular Disease | ICD10 | I77.1 | Chronic Pulmonary Disease | ICD10 | J44 |
| Periphral Vascular Disease | ICD10 | I79.0 | Chronic Pulmonary Disease | ICD10 | J45 |
| Periphral Vascular Disease | ICD10 | I79.2 | Chronic Pulmonary Disease | ICD10 | J46 |
| Periphral Vascular Disease | ICD10 | K55.1 | Chronic Pulmonary Disease | ICD10 | J47 |
| Periphral Vascular Disease | ICD10 | K55.8 | Chronic Pulmonary Disease | ICD10 | J60 |
| Periphral Vascular Disease | ICD10 | K55.9 | Chronic Pulmonary Disease | ICD10 | J61 |
| Periphral Vascular Disease | ICD10 | Z95.8 | Chronic Pulmonary Disease | ICD10 | J62 |
| Periphral Vascular Disease | ICD10 | Z95.9 | Chronic Pulmonary Disease | ICD10 | J63 |
| Connective Tissue Disease-Rheumatic Disease | ICD10 | M05 | Chronic Pulmonary Disease | ICD10 | J64 |
| Connective Tissue Disease-Rheumatic Disease | ICD10 | M32 | Chronic Pulmonary Disease | ICD10 | J65 |
| Connective Tissue Disease-Rheumatic Disease | ICD10 | M33 | Chronic Pulmonary Disease | ICD10 | J66 |
| Connective Tissue Disease-Rheumatic Disease | ICD10 | M34 | Chronic Pulmonary Disease | ICD10 | J67.I278 |
| Connective Tissue Disease-Rheumatic Disease | ICD10 | M06 | Chronic Pulmonary Disease | ICD10 | I27.9 |
| Connective Tissue Disease-Rheumatic Disease | ICD10 | M31.5 | Chronic Pulmonary Disease | ICD10 | J68.4 |
| Connective Tissue Disease-Rheumatic Disease | ICD10 | M35.1 | Chronic Pulmonary Disease | ICD10 | J70.1 |
| Connective Tissue Disease-Rheumatic Disease | ICD10 | M35.3 | Chronic Pulmonary Disease | ICD10 | J70.3 |
| Connective Tissue Disease-Rheumatic Disease | ICD10 | M36.0 |  |  |  |
| Peptic Ulcer Disease | ICD10 | K25 |  |  |  |
| Peptic Ulcer Disease | ICD10 | K26 |  |  |  |
| Peptic Ulcer Disease | ICD10 | K27 |  |  |  |
| Peptic Ulcer Disease | ICD10 | K28 |  |  |  |

**eTable 2. Health Utilization Survey Question Categorizations**

| **Category Name** | **Question found in Survey** | **Number of respondents** |
| --- | --- | --- |
| Mutual Relationship with Provider | How often did your doctors or health care providers ask for your opinions or beliefs about your medical care or treatment? For example, what kind of tests, procedures, or medications you prefer. | 7,498 |
|  | How often did your doctors or health care providers tell or give you information about your health and health care that was easy to understand? | 7,585 |
|  | How often were you treated with respect by your doctors or health care providers? | 7,604 |
| Delayed Care Due to Lack of Diversity | How often have you either delayed or not gone to see doctors or health care providers because they were different from you in any of these ways? | 7,528 |
| Financial Barrier | If you get sick or have an accident, how worried are you that you will be able to pay your medical bills? Are you very worried, somewhat worried, or not at all worried? | 7,572 |
|  | DURING THE PAST 12 MONTHS, were any of the following true for you: |  |
|  | You used alternative therapies to save money | 6,973 |
|  | You bought prescription drugs from another country to save money | 7,054 |
|  | You delayed filling a prescription to save money | 7,502 |
|  | You asked your doctor for a lower cost medication to save money | 7,213 |
|  | You skipped medication doses to save money | 7,751 |
|  | You took less medicine to save money | 7,786 |
| Provider Inaccessibility | Is there a place that you USUALLY go to when you are sick or need advice about your health? | 7,633 |
|  | About how long has it been since you last saw or talked to a doctor or other health care provider about your own health? | 7,601 |
| Insurance Barrier | DURING THE PAST 12 MONTHS, were you told by a health care provider or doctor’s office that they did not accept your health care coverage? | 7,689 |
|  | Have you delayed getting care for any of the following reasons in the PAST 12 MONTHS: |  |
|  | Couldn’t afford the copay. | 6,534 |
|  | Your deductible was too high/or could not afford the deductible. | 6,470 |
|  | You had to pay out of pocket for some or all of the procedure. | 6,474 |
|  | In regard to your health insurance or health care coverage, how does it compare to a year ago? Is it better, worse, or about the same? | 7,631 |
| Patient Anxiety | Have you delayed getting care for any of  the following reasons in the PAST 12 MONTHS:  You were nervous about seeing your healthcare provider. | 7,252 |
| Rural Barrier | Have you delayed getting care for any of the following reasons in the PAST 12 MONTHS: You live in a rural area where distance to the health care provider is too far. | 7,501 |
| Social Dependency | Have you delayed getting care for any of the following reasons in the PAST 12 MONTHS: Couldn’t get childcare. | 6,549 |
| Transportation Barrier | Have you delayed getting care for any of the following reasons in the PAST 12 MONTHS: Didn’t have transportation. | 7,628 |
| Employment Barrier | Have you delayed getting care for any of the following reasons in the PAST 12 MONTHS: Couldn’t get time off work. | 6,952 |

**eTable 3:** **Bivariate Analysis of the Survey Cohort According to Percent Days Covered (PDC) of Statin Prescription**

|  | Survey Cohort | | |
| --- | --- | --- | --- |
|  | **PDC < 0.80**  **N= 4734** | **PDC => 0.80**  **N= 3052** | **Odds Ratio**  **(95% CI)** |
| Age (mean (SD)) | 64.33 (11.20) | 65.85 (10.09) | 1.01 (1.01, 1.02) |
| Sex at Birth – Female (ref: Male) (%) | 2416 (51.0) | 1493 (48.9) | 0.92 (0.84, 1.01)) |
| Race (%) |  |  |  |
| Non-Hispanic Black | 650 (13.7) | 407 (13.3) | 0.91 (0.8, 1.04) |
| Hispanic | 330 (7.0) | 62 (2.0) | 0.27 (0.21, 0.36) |
| Non-Hispanic White | 3754 (79.3) | 2583 (84.6) | Reference |
| Insurance (%) |  |  |  |
| Medicaid | 1826 (38.6) | 1239 (40.6) | 1.11 (0.96, 1.29) |
| Medicare | 722 (15.3) | 455 (14.9) | 1.2 (1.07, 1.34) |
| Other | 290 (6.1) | 169 (5.5) | 1.03 (0.83, 1.27) |
| Private | 1330 (28.1) | 754 (24.7) |  |
| VA | 566 (12.0) | 435 (14.3) | 1.36 (1.16, 1.58) |
| Missing | - | - | - |
| Education (%) |  |  |  |
| Less than secondary | 77 (1.6) | 29 (1.0) | 0.58 (0.37, 0.88) |
| Secondary | 821 (17.3) | 527 (17.3) | 0.99 (0.87, 1.11) |
| Post-Secondary | 3836 (81.0) | 2496 (81.8) | Reference |
| Missing | - | - | - |
| Income (%) |  |  |  |
| Low <$ 35K | 1564 (33.0) | 980 (32.1) | 1.02 (0.91, 1.14) |
| Middle $35K – $75K | 1410 (29.8) | 992 (32.5) | 1.15 (1.03, 1.28) |
| High >$ 75k | 1760 (37.2) | 1080 (35.4) | Reference |
| Missing |  |  |  |
| Foreign Born (%) | 311 (6.6) | 91 (3.0) | 0.44 (0.34, 0.55) |
| Not Employed (%) | 3037 (64.2) | 2103 (68.9) | 1.24 (1.12, 1.36) |
| Marital Status (%) |  |  |  |
| Married/Widowed | 2793 (59.0) | 1805 (59.1) | 1 (0.87, 1.15) |
| Separated/Divorce | 1356 (28.6) | 869 (28.5) | 0.99 (0.85, 1.16) |
| Single | 585 (12.4) | 378 (12.4) | Reference |
| Missing |  |  |  |
| Smoking (%) |  |  |  |
| Current | 491 (10.4) | 280 (9.2) | 0.91 (0.77, 1.06) |
| Former | 1798 (38.0) | 1234 (40.4) | 1.09 (0.99, 1.2) |
| Never | 2445 (51.6) | 1538 (50.4) |  |
| Missing | - | - | - |
| CCI High (%) | 3543 (74.8) | 2446 (80.1) | 1.36 (1.22, 1.52) |
| Serum Creatinine (mean (SD)) | 2.5 (3.6) | 1.9 (2.9) | 0.94 (0.93, 0.96) |
| SBP (mean (SD)) | 131.0 (16.6) | 131.2 (16.4) | 1 (1,1) |
| HDL (mean (SD)) | 38.0 (16.4) | 38.5 (14.9 | 1 (1, 1) |
| LDL (mean (SD)) | 90.7 (36.4) | 81.0 (30.7) | 0.99 (0.99,0.99) |
| Total Chol (mean(SD)) | 177.5 (45.0) | 167.6 (38.9) | 0.99 (0.99, 1) |
| DBP (mean (SD)) | 77.7 (10.3) | 76.9 (10.0) | 0.99 (0.99,0.99) |
| BMI (mean (SD)) | 31.7 (7.2) | 31.8 (7.0) | 0.99 (0.99,0.99) |
| MI (%) | 474 (10.0) | 343 (11.2) | 1.14 (0.98, 1.32) |
| CHF (%) | 616 (13.0) | 428 (14.0) | 1.09 (0.95, 1.24) |
| Liver Disease (%) | 772 (16.3) | 518 (17.0) | 1.05 (0.93, 1.18) |
| Any Diabetes (%) | 1630 (34.4) | 1171 (38.4) | 1.19 (1.08, 1.3) |

*Continuous variables are reported as means with standard deviations, and categorical variables are reported as percentages. Unadjusted odds ratios (95%CI) are reported for each variable.*

^†^ Other race category includes American Indian/Alaskan native, Middle Eastern, Asian, Native Hawaiian/ Pacific Islander.

Abbreviations: PDC: Percent Days Covered, CI: Confidence Interval, SD: Standard Deviation, CCI: Charlson Comorbidity Index, SBP: Systolic Blood Pressure, HDL: High Density Lipoprotein, LDL: Low Density Lipoprotein, Chol: Cholesterol, DBP: Diastolic Blood Pressure, BMI: Body Mass Index, MI: Myocardial Infarction, CHF: Congestive Heart Failure
